# Online group-based Acceptance and Commitment Therapy for stroke survivors: a study of fidelity of delivery within the Wellbeing After Stroke study

**DOI:** 10.1101/2024.09.05.24313107

**Authors:** Hannah Foote, Audrey Bowen, Sarah Cotterill, Emma Patchwood

## Abstract

**Objectives:** To explore whether practitioners can deliver the Acceptance and Commitment Therapy-based Wellbeing After Stroke intervention with fidelity to both the clinical protocol and the Acceptance and Commitment therapy model.

**Design:** Observational fidelity study, embedded within the Wellbeing After Stroke study. Setting: online groups. UK.

**Participants:** Practitioners employed by the Stroke Association, trained to deliver the intervention.

**Measures:** 1) a bespoke Wellbeing After Stroke fidelity tool to assess fidelity to dose, duration and content of intervention sessions, self-completed by practitioners and a sub-set completed by researchers based on video recordings. We calculated inter-rater reliability of researchers and practitioners. 2) Acceptance and Commitment Therapy-Fidelity Measure to assess fidelity to the Acceptance and Commitment Therapy model, completed by researchers on the sub-set of recorded sessions.

**Results:** Seven practitioners delivered the Wellbeing After Stroke intervention to three groups of stroke survivors. The planned dose of the intervention was delivered, with duration slightly longer than planned. Practitioners delivered the intervention with high fidelity to protocol: 92–100% of content delivered, as measured by the Wellbeing After Stroke fidelity tool, once reliability was established. Some practitioners delivered the intervention with fidelity to the Acceptance and Commitment Therapy model.

**Conclusions:** Trained and supervised practitioners can deliver an online, group Acceptance and Commitment Therapy-based intervention to stroke survivors with high fidelity to protocol. Improving training may increase consistency with the Acceptance and Commitment Therapy model. The ACT-Fidelity Measure can be used to measure consistency of delivery of protocolised, group interventions, but adaptations would increase suitability to context.

## Introduction

Stroke survivors frequently experience mental health difficulties and many services cannot fully meet their needs^1–3^. Psychological interventions for those with mild-to-moderate needs can be delivered by non-specialist staff, with appropriate training and supervision ^4,5^. One such intervention is Acceptance and Commitment Therapy ^6^, a trans-diagnostic, third-wave, cognitive behavioural therapy, with growing evidence for its use for psychological difficulties post-stroke ^7–11^.

The Wellbeing After Stroke study developed and demonstrated the feasibility of delivering a nine-week, online, protocolised intervention, informed by Acceptance and Commitment Therapy, to groups of stroke survivors ^12^. An adjunct training programme upskilled practitioners without previous experience of this therapy, to deliver the intervention under the supervision of a clinical neuropsychologist. A detailed intervention and training description is provided in the supplemental materials of the feasibility paper^12^. The feasibility paper ^12^ did not robustly report intervention fidelity: a multi-dimensional construct referring to the extent to which an intervention is delivered and received as intended and which can affect outcomes ^13,14^.

The present paper focuses on fidelity, exploring both **what** the practitioners delivered, i.e. examining whether they delivered the components of the clinical protocol; and **how** the practitioners delivered the intervention, i.e. examining whether they had fidelity to the therapeutic model.

## Research objectives were to explore

1. the reliability of practitioners self-monitoring their fidelity.
2. whether the intervention was delivered to protocol and reasons for protocol deviations.
3. whether the Wellbeing After Stroke intervention was delivered in a manner consistent with the Acceptance and Commitment Therapy model.

## Methods

Research ethics approval was secured from the University of Manchester (ref 2021-11134-18220).

Participants were those already recruited and trained as lead and support practitioners in the broader Wellbeing After Stroke study ^12^. The eligibility criteria were:

- Employed as frontline practitioners by Stroke Association (a UK national charity specialising in stroke) for at least 6 months
- Capacity and willingness to participate with clearance and support from their line manager
- Experience and knowledge of facilitating groups of stroke survivors Two tools were used for exploring research objectives:

**1. Wellbeing After Stroke fidelity tool** (see supplemental materials): a bespoke tool developed by authors to monitor delivery of components of the clinical protocol. Data collected:

- Whether practitioners delivered each content component of each session (112 components across all nine sessions). Scoring was a binary yes/no rating, with ‘yes’ indicating that they had fully delivered a component and ‘no’ indicating that they had partially or not delivered a component
- Date, time and attendance at each session, and practitioner judgement on session length (‘too short’, ‘about right’ or ‘too long’)

- Practitioners’ reasons for not fully delivering any component (free-text question)
- Any other comments on the session (free-text question).

**2. Acceptance and Commitment Therapy – Fidelity Measure** ^15^. A published tool, designed to explore practitioner fidelity with the therapeutic model. The ACT-Fidelity Measure explores four areas of Acceptance and Commitment Therapy delivery: Therapist Stance, Open Response Style, Aware Response Style and Engaged Response style. Each area is scored from 0–9 for consistency and 0–9 for inconsistency, giving a total consistency score and a total inconsistency score, each from 0–36. Researchers (HF and EP) completed training on use of the tool and score calibration prior to use (training plans agreed with Acceptance and Commitment Therapy-Fidelity Measure authors Lucy O’Neill and Christopher Graham, via emails exchanged in February 2021).

The Wellbeing After Stroke intervention consisted of nine weekly sessions ^12^. Three intervention groups ran (Groups A, B and C), each delivered by two practitioners (one lead and one support). All intervention sessions were video-recorded, and researchers used these recordings to collect data from a sub-set of nine sessions using both fidelity tools. Due to time and resource limitations this sample included sessions from Groups A and B only. In addition, to test self-completion of the new Wellbeing After Stroke fidelity tool, lead and support practitioners were trained on its use and asked to independently complete the tool immediately following each session (to aid recall), and email this to the research team within 24 hours of each session finishing. They were prompted by an email reminder within this time period. This was done for all sessions for all three groups. Table 1 summarises how the two fidelity tools were used to address each research objective.

**Table 1:**
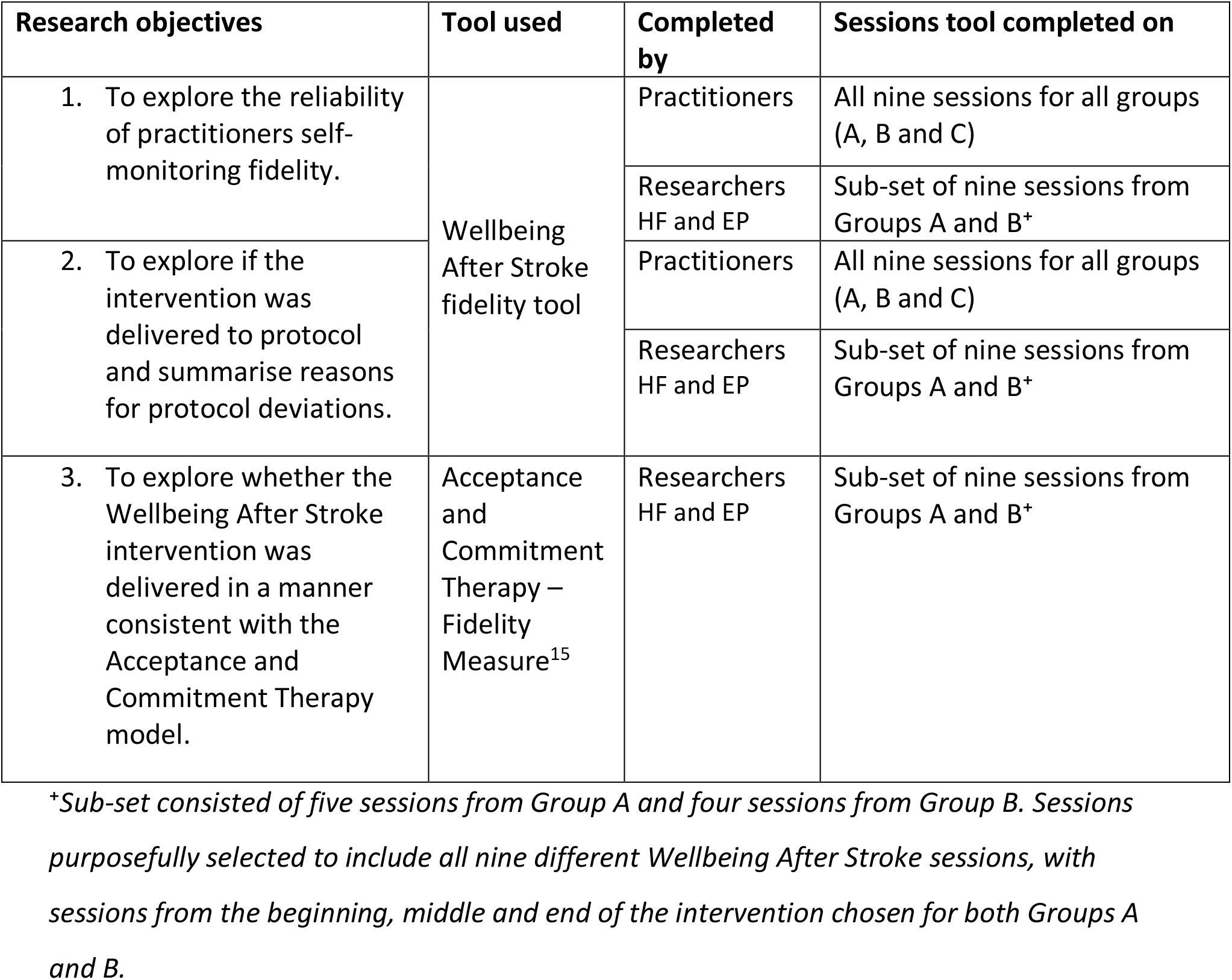
Summary of how the tools were used to answer each research objective.

To explore the reliability of practitioners self-monitoring fidelity, inter-rater reliability on completion of the Wellbeing After Stroke fidelity tool was calculated, first between the lead and support practitioners, and secondly, between the researcher and lead practitioner, calculated using the Prevalence-Adjusted and Bias-Adjusted Kappa (PABAK) statistic ^16^. Sufficient reliability was set *a priori* as being at least good agreement (kappa between 0.61 and 0.8) ^17^.

To explore if the intervention was delivered to protocol and summarise reasons for protocol deviations, data from the Wellbeing After Stroke fidelity tool were analysed and summarised as follows:

- The percentage of content items delivered (per group and totalled for all three groups)
- Frequency and duration of sessions, including summarising practitioners’ judgement on session length, and the total percentage of sessions attended by stroke survivors across all three groups
- For the free-text questions both the lead and support practitioners’ data were analysed to add insight to the quantitative data

Data analysis used the researcher ratings available for the sub-set of nine sessions, and, if inter-rater reliability on the tool had been established, used the lead practitioner ratings for other sessions.

To explore whether the Wellbeing After Stroke intervention was delivered in a manner consistent with the therapeutic model, the Acceptance and Commitment Therapy-Fidelity Measure^15^ was rated by the researchers observing videos of the sub-set of nine intervention sessions (as per Table 1). Published guidelines for this measure do not provide cut-off scores as to what are adequate level of in/consistency or guidance on how to combine the scores. Therefore no *a priori* level to be reached was set. Scores were calculated for each group: both overall (total and mean) and per area.

## Results

Eight practitioners were trained and recruited as part of the broader Wellbeing After Stroke study, and seven of these delivered at least one intervention session. All seven practitioners contributed data to this fidelity study. Four were lead practitioners, and three were support. In addition to the eligibility criteria, the lead practitioners all had a level four counselling qualification. All were female, with a mean age of 51.6 years (SD: 8.03). The mean number of years working for the Stroke Association was 5.1 years (range 1–15 years).

For objective one, the inter-rater reliability of the use of Wellbeing After Stroke fidelity tool was calculated and interpreted in accordance with Altman ^17^. Agreement in Group A was ‘very good’ for lead and support practitioners (0.89 PABAK; 95% CIs 0.81 to 0.98), and lead and research (0.90 PABAK; 95% CIs 0.79 to 1). Agreement in group C for lead and support was ‘perfect’ (i.e. ratings did not vary) and not calculated for lead and researchers (as Group C ratings were not included in the researcher sub-set). Agreement in Group B was ‘good’ for lead and support practitioners (0.80 PABAK; 95% Cis 0.69 to 0.01) and lead and researcher (0.65 PABAK; 95% CIs 0.44 to 0.77). Practitioners were able to reliably complete with at least ‘good’ agreement in all cases, and therefore analyses of Wellbeing After Stroke fidelity tool data for objective two could be completed as planned.

For objective two, the Wellbeing After Stroke fidelity tool showed that Group A delivered 92% (103/112) of all protocolised components, Group B delivered 96% (108/112) and Group C delivered 100% (112/112). In total, 96% (323/336) of all protocol components were delivered across the three groups.

Free-text practitioner comments stated that both the group supervision sessions (held weekly with a clinical neuropsychologist) and having two practitioners per group supported successful delivery of the groups. Technological issues were occasionally reported as negatively impacting (but not preventing) delivery of components, e.g. “my internet dropped at the end of the session and [support practitioner] took over” [ID05]. Suggestions to improve the clinical protocol were given, such as moving the order of activities to better suit timings.

All sessions happened weekly as planned, with 98% attendance across the three groups (two stroke survivors missed one session each, one due to being away and one due to a power outage). There were three occasions of practitioner absence, but these were known in advance and so the sessions were covered by a different practitioner.

The sessions were all planned to be 120 minutes duration. Overall, the mean length of delivered sessions was 128 minutes (min: 60 minutes, max: 150 minutes), but actual duration varied according to session number (from 95-145 minutes) (see Table 2). The mean session length was shorter than planned for Sessions One and Three, and longer than planned in all other sessions.

**Table 2:**
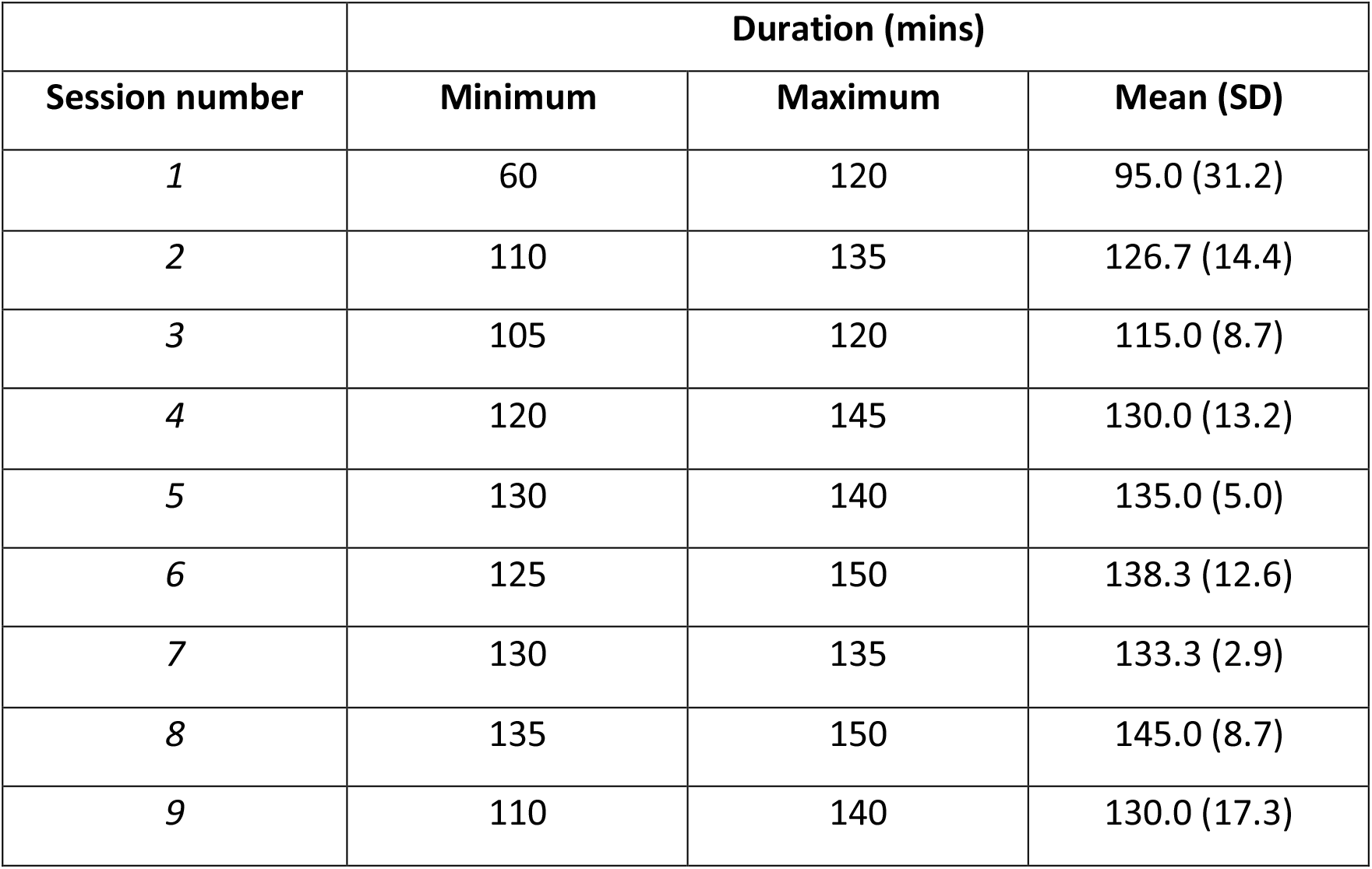
Actual intervention session duration.

Of the twenty-seven lead practitioner ratings of duration (nine sessions for each of the three groups), twenty sessions were rated as ‘about right’, 6 as ‘too short’ and 1 (Group 3, session 1) as ‘too long’. The free-text responses give some insight into why practitioners had rated the time available for a session as ‘about right’ despite a longer duration time than anticipated. For example, “we overran by half an hour but there was quite a lot of reflecting and talking. I don’t think the session is too short.” [ID01].

When the practitioners did rate a session as ‘too short’ comments indicate that practitioners were concerned that components were not sufficiently explained or discussed, e.g. “we rushed through the home practice for next week” [ID07] and “more time could be given to such an emotive topic” [ID05]. Lack of time was the most common reason the practitioners gave for partially/not delivering an intervention component, indicating that the practitioners were mitigating for even longer session durations. For example, “I was mindful of the time and although I invited feedback and checked for understanding, I felt that I skipped over this quite quickly.” [ID05].

Other comments made by the practitioners related to whether the intervention was received with fidelity by the stroke survivors. Most of these comments were positive, for example, “[stroke survivors] engaged well in the session both with the facilitators [practitioners] and each other” [ID01], and reported the stroke survivors engaging in the homework, e.g. “all [stroke survivors] are engaging well in the home practice” [ID07].

Conversely, some comments noted that stroke survivors struggled with understanding “one [stroke survivor] could not grasp the concept” [ID01] and lack of homework engagement “I don’t feel that many [stroke survivors] are actively engaging with much of the home practice” [ID05]. Post-stroke difficulties presented a barrier to engagement on occasion, e.g. one stroke survivor “could physically write but found it difficult to organise [their] thoughts and language to record” [ID06].

For objective three, Table 3 shows researcher scores on the Acceptance and Commitment Therapy-Fidelity Measure^15^ based on video observations. The total in/consistency scores suggest that both Group A and B practitioners were more Acceptance and Commitment Therapy consistent than they were inconsistent. However, there is a different profile of scores, with Group A having both higher consistency and lower inconsistency scores than Group B. Group B practitioners were observed to stick less closely to the scripts given in the protocol and scored as less consistent with Acceptance and Commitment Therapy during the unscripted sections of the protocol.

**Table 3:** Acceptance and Commitment Therapy in/consistency scores for Groups A and B.

| **Group A consistency and inconsistency scores (across 5 rated sessions)** |  |  |  |
| --- | --- | --- | --- |
| **Subscales - consistent** | **Mean** | **Subscales - inconsistent** | **Mean** |
| Therapist stance (0–9) | 5 | Therapist stance (0–9) | 0.2 |
| Open (0–9) | 5.4 | Open (0–9) | 1.6 |
| Aware (0–9) | 5.6 | Aware (0–9) | 0 |
| Engaged (0–9) | 5.2 | Engaged (0–9) | 0.2 |
| **Total consistency (0–36)** | **21.2** | **Total inconsistency (0–36)** | **2** |
| **Group B consistency and inconsistency scores (across 4 rated sessions)** |  |  |  |
| **Subscales - consistent** | **Mean** | **Subscales - inconsistent** | **Mean** |
| Therapist stance (0–9) | 3.9 | Therapist stance (0–9) | 2.8 |
| Open (0–9) | 1.8 | Open (0–9) | 3.8 |
| Aware (0–9) | 3.8 | Aware (0–9) | 1.5 |
| Engaged (0–9) | 4.5 | Engaged (0–9) | 2 |
| **Total consistency (0–36)** | **14** | **Total inconsistency (0–36)** | **10.1** |

The Group A results are more Acceptance and Commitment Therapy consistent than inconsistent across all subscales, as are the Group B results, with the exception of the ‘Open’ subscale. This subscale also has the highest inconsistency score in Group A results.

## Discussion

Practitioners, trained and supervised by a clinical neuropsychologist, were able to deliver an online, group Acceptance and Commitment Therapy-informed intervention to stroke survivors according to protocol and were reliable in self-monitoring fidelity. Intervention dose was as planned and attendance was high. Almost all intervention components were delivered, but most sessions were slightly longer than planned. Some practitioners delivered the intervention with high fidelity to the therapeutic model. The Acceptance and Commitment Therapy – Fidelity Measure^15^ was found to be useable in a novel context (a protocolised online, group intervention), but adaptations would improve fit to context.

Practitioners had high fidelity to the Wellbeing After Stroke study protocol (covering 92-100% of the protocol), meeting the 80% threshold recommended by Borrelli ^18^ and comparing favourably to previous findings from a single group intervention (94%) ^9^ and four individual cases (60%, 80%, 80%, 80%) ^10^. To our knowledge, the Acceptance and Commitment Therapy-Fidelity Measure^15^ has not previously been used for a post-stroke intervention. The guidelines for this measure do not provide cut-off scores as to what is adequate consistency with the therapeutic model, however, a recently published trial protocol of Acceptance and Commitment Therapy for a multiple sclerosis population proposed cut-off scores for low/high fidelity^19^. When applying these cut-offs to the present study’s results, practitioners in Group B had low fidelity and practitioners in Group A had high fidelity, indicating that the Wellbeing After Stroke training may not be sufficient to enable all practitioners to deliver the intervention with adequate fidelity to the therapeutic model. However, these proposed cut-offs were designed for use with psychologists rather than practitioners, and a higher level of consistency may be expected for the former group.

A strength of our study is the use of two tools. The published Acceptance and Commitment Therapy-Fidelity Measure^15^ was developed by experts, however it requires further psychometric evaluation and does not have guidance on score interpretation. The measure was not designed for use with a protocolised group intervention and certain items were less suited to this context. Furthermore, in the Wellbeing After Stroke protocol, certain sessions had a specific focus, and so opportunities for scoring on all aspects of the measure were limited. Tailoring the measure to different intervention contexts may be beneficial and has been done in studies with different populations by adapting scoring to reflect didactic delivery^20^ and weighting scores differently per session to reflect the session’s focus^21^.

The Wellbeing After stroke fidelity tool was developed and used for the first time in this study. The tool is self-completed by practitioners, which can be less resource-intensive than external rating and may be practicable for real-world implementation. However, self-completion tools are subjective and can lead to response bias. To mitigate against this, researchers rated a sub-sample of the sessions and reliability was found to be good.

Researcher ratings of the practitioners were collected from two of the three groups, due to limited resources. Practitioners were delivering the intervention for the first time in these groups and so we do not know if levels of fidelity will change over time. Lead practitioners all had a counselling qualification (although this was not an eligibility criteria), which may have impacted on their levels of fidelity. This study focused primarily on fidelity of delivery, with only incidental data as to fidelity of receiving the intervention. The Wellbeing After Stroke intervention was feasible to deliver^12^ and acceptable to stroke survivors ^22^.

In future research studies (Wellbeing After Stroke-2 began October 2023^23^), fidelity to the duration of sessions will be explored. Duration may reduce over time as practitioners become more experienced. Alternatively, the training may require increased focus on time-management, or the intervention may require a reduction in content (ensuring no essential components are removed) or an increase in session length (with consideration of burden).

Further research on the Acceptance and Commitment Therapy – Fidelity Measure^15^, with a larger sample of practitioners, could establish cut-off scores for the delivery of a protocolised intervention with adequate fidelity to the therapeutic model, and explore whether adaptations to the measure would be beneficial. Further research to optimise the Wellbeing After Stroke training may support the achievement of an adequate level of fidelity to the model, particularly in having an ‘open response style’. Practitioners self-rating Acceptance and Commitment Therapy-consistency could lead to increased self-monitoring, and potentially increase fidelity. Future research exploring whether interventions are *received* with fidelity, may enable intervention optimisation and support implementation^24^.

In conclusion, this study suggests that it is possible for practitioners to deliver an online, group Acceptance and Commitment Therapy intervention to stroke survivors with high fidelity to protocol, following a brief training course and with weekly supervision from a clinical neuropsychologist. However, improvements to training may improve their fidelity to the Acceptance and Commitment Therapy model. This study indicates that it is possible to use the Acceptance and Commitment Therapy-Fidelity Measure^15^ for group, protocolised interventions, but that adaptations may support better fit to context. Further research exploring the level of fidelity to the Acceptance and Commitment Therapy model required for successful delivery of such groups would be beneficial. This study strengthens the evidence base for high fidelity of delivery to protocolised Acceptance and Commitment Therapy interventions within acquired brain injury populations.

## Supporting information

Supplemental Materials: The Wellbeing After Stroke (WAterS) fidelity tool

## Data Availability

All data produced in the present study are available upon reasonable request to the authors

## Availability of Data

All data referred to in this manuscript can be requested via the corresponding author. All requests will be dealt with on a case-by-case basis.

