## Supplemental Materials: The Wellbeing After Stroke (WAterS) fidelity tool for "Online group-based Acceptance and Commitment Therapy for stroke survivors: a study of fidelity of delivery within the Wellbeing After Stroke study"

**Supplemental material: WAterS fidelity tool**

WAterS fidelity checklist session 1 - Coming together after stroke

*Please remember:*

- *Complete this ASAP following the session*
- *Fill this out honestly. Remember this is for our learning, not to penalise you*
- *Yes = component fully delivered as per protocol*
- *No = component not delivered or partially delivered*

| Date: | Start time: | End time: |
|---|---|---|
| Name of facilitator/s: | | |
| Number of participants: | | |

| Component | | Delivered as planned? | |
|---|---|---|---|
| Beginning the session (50 mins) | | | |
| A | Introduction to facilitator and participants | No☐ | Yes☐ |
| B | Outline of session one | No☐ | Yes☐ |
| C | Introduction to the client handbook [handbook] | No☐ | Yes☐ |
| D | Introduce ACT and the aims of the course | No☐ | Yes☐ |
| E | Outline the structure of the programme [handbook] | No☐ | Yes☐ |
| Group rules (15 mins) | | | |
| F | Core rules [handbook] | No☐ | Yes☐ |
| G | Adequate opportunity to discuss any additional rules | No☐ | Yes☐ |
| Ending the session (20 mins) | | | |
| H | Summary of the session | No☐ | Yes☐ |
| I | Adequate opportunity for reflections & comments | No☐ | Yes☐ |
| J | Home practice [handbook] | No☐ | Yes☐ |

<table>
<tr><td colspan="4">If you’ve answered ‘No’ for any components, please give your reasoning (use the component’s letter to identify which component/s you are referring to):<br><br>Please type as much as you like</td></tr>
<tr><td>2 hours (approximately) to deliver all the session components was:</td><td>Too short☐</td><td>About right☐</td><td>Too long☐</td></tr>
<tr><td colspan="4">Any other comments?<br><br>Please type as much as you like</td></tr>
</table>

WAterS fidelity checklist session 2 - How can I live better after stroke?

*Please remember:*

- *Complete this ASAP following the session*
- *Fill this out honestly. Remember this is for our learning, not to penalise you*
- *Yes = component fully delivered as per protocol*
- *No = component not delivered or partially delivered*

| Date: | Start time: | End time: |
|---|---|---|
| Name of facilitator/s: | | |
| Number of participants: | | |

| Component | | Delivered as planned? | |
|---|---|---|---|
| Beginning the session (40 mins) | | | |
| A | Re-introductions to facilitator and participants | No☐ | Yes☐ |
| B | Recap on session 1 [handbook] | No☐ | Yes☐ |
| C | Outline session 2 | No☐ | Yes☐ |
| D | Home practice feedback | No☐ | Yes☐ |
| Beyond barriers to group access: Making room for forgetting (15 mins) | | | |
| E | Making room for forgetting [slides] | No☐ | Yes☐ |
| F | Adequate opportunity for reflections & comments | No☐ | Yes☐ |
| What do you value? Clarification and construction exercise (30 mins) | | | |
| G | What are values? [slides] | No☐ | Yes☐ |
| H | Adequate opportunity for reflections & comments | No☐ | Yes☐ |
| I | Activity – what are my values? [handbook] | No☐ | Yes☐ |
| J | Adequate opportunity for reflections & comments | No☐ | Yes☐ |
| K | Choice points [slides] | No☐ | Yes☐ |

| | | | |
|---|---|---|---|
| L | Adequate opportunity for reflections & comments | No☐ | Yes☐ |
| Ending the session (20 mins) | | | |
| M | Summary of the session | No☐ | Yes☐ |
| N | Adequate opportunity for reflections & comments | No☐ | Yes☐ |
| O | Home practice [handbook] | No☐ | Yes☐ |
| If you've answered 'No' for any components, please give your reasoning (use the component's letter to identify which component/s you are referring to):<br><br>*Please type as much as you like* | | | |

| | | | |
|---|---|---|---|
| 2 hours (approximately) to deliver all the session components was: | Too short☐ | About right☐ | Too long☐ |
| Any other comments?<br><br>*Please type as much as you like* | | | |

WAterS fidelity checklist session 3 – Noticing more

*Please remember:*

- *Complete this ASAP following the session*
- *Fill this out honestly. Remember this is for our learning, not to penalise you*
- *Yes = component fully delivered as per protocol*
- *No = component not delivered or partially delivered*

| Date: | Start time: | End time: |
|---|---|---|
| Name of facilitator/s: | | |
| Number of participants: | | |

| Component | | Delivered as planned? | |
|---|---|---|---|
| Beginning the session (30 mins) | | | |
| A | Re-cap on session 2 | No☐ | Yes☐ |
| B | Outline session 3 | No☐ | Yes☐ |
| C | Home practice feedback | No☐ | Yes☐ |
| Beyond barriers to access - Allowing fatigue (15 mins) | | | |
| D | Allowing fatigue [slides] | No☐ | Yes☐ |
| E | Adequate opportunity for reflections & comments | No☐ | Yes☐ |
| Mindful noticing (35 mins) | | | |
| F | Introduction to mindful noticing [slides] | No☐ | Yes☐ |
| G | Lead the guided noticing of an object exercise [object] | No☐ | Yes☐ |
| H | Undertake an enquiry, supporting the group to share what they noticed [handbook] | No☐ | Yes☐ |
| Ending the session (20 mins) | | | |
| I | Summary of the session | No☐ | Yes☐ |
| J | Adequate opportunity for reflections & comments | No☐ | Yes☐ |
| K | Home practice [handbook] | No☐ | Yes☐ |

| | | | |
|---|---|---|---|
| If you’ve answered ‘No’ for any components, please give your reasoning (use the component’s letter to identify which component/s you are referring to):<br><br>*Please type as much as you like* | | | |
| 2 hours (approximately) to deliver all the session components was: | Too short☐ | About right☐ | Too long☐ |
| Any other comments?<br><br>*Please type as much as you like* | | | |

WAterS fidelity checklist Session 4 – Becoming more present

*Please remember:*

- *Complete this ASAP following the session*
- *Fill this out honestly. Remember this is for our learning, not to penalise you*
- *Yes = component fully delivered as per protocol*
- *No = component not delivered or partially delivered*

| Date: | Start time: | End time: |
|---|---|---|
| Name of facilitator/s: | | |
| Number of participants: | | |

| Component | | Delivered as planned? | |
|---|---|---|---|
| Beginning the session (35 mins) | | | |
| A | Re-cap on session 3 | No ☐ | Yes ☐ |
| B | Outline session 4 | No ☐ | Yes ☐ |
| C | Home practice feedback | No ☐ | Yes ☐ |
| Beyond barriers to group access: Opening up to word-finding problems (15 mins) | | | |
| D | Opening up to word-finding problems [slides] | No ☐ | Yes ☐ |
| E | Adequate opportunity for reflections & comments | No ☐ | Yes ☐ |
| Mindful noticing (35 mins) | | | |
| F | Introduction to guided mindful noticing practices | No ☐ | Yes ☐ |
| G | Guided noticing of the body [audio] | No ☐ | Yes ☐ |
| H | Undertake an enquiry, guided by example questions | No ☐ | Yes ☐ |
| Ending the session (20 mins) | | | |
| I | Summary of the session | No ☐ | Yes ☐ |
| J | Adequate opportunity for reflections & comments | No ☐ | Yes ☐ |

| | | | |
|---|---|---|---|
| K | Home practice [handbook] | No ☐ | Yes ☐ |
| If you’ve answered ‘No’ for any components, please give your reasoning (use the component’s letter to identify which component/s you are referring to):<br><br>*Please type as much as you like* | | | |

| | | | |
|---|---|---|---|
| 2 hours (approximately) to deliver all the session components was: | Too short☐ | About right☐ | Too long☐ |
| Any other comments?<br><br>*Please type as much as you like* | | | |

WAterS fidelity checklist Session 5 – Common feelings after stroke

*Please remember:*

- *Complete this ASAP following the session*
- *Fill this out honestly. Remember this is for our learning, not to penalise you*
- *Yes = component fully delivered as per protocol*
- *No = component not delivered or partially delivered*

| Date: | Start time: | End time: |
|---|---|---|
| Name of facilitator/s: | | |
| Number of participants: | | |

| Component | | Delivered as planned? | |
|---|---|---|---|
| Beginning the session (30 mins) | | | |
| A | Re-cap on session 4 | No ☐ | Yes ☐ |
| B | Outline session 5 | No ☐ | Yes ☐ |
| C | Home practice feedback | No ☐ | Yes ☐ |
| Guided noticing of the body with enquiry (30 mins) | | | |
| D | Guided noticing of the body [audio] | No ☐ | Yes ☐ |
| E | Undertake an enquiry – guided by example questions | No ☐ | Yes ☐ |
| Common feelings after stroke (30 mins) | | | |
| F | Common feelings after stroke [slides] | No ☐ | Yes ☐ |
| G | Activity – choose and notice an emotion [handbook] | No ☐ | Yes ☐ |
| H | Adequate opportunity for reflections & comments | No ☐ | Yes ☐ |
| Ending the session (20 mins) | | | |
| I | Summary of the session | No ☐ | Yes ☐ |
| J | Adequate opportunity for reflections & comments | No ☐ | Yes ☐ |
| K | Home practice [handbook] | No ☐ | Yes ☐ |

<table>
<tr><td colspan="4">If you’ve answered ‘No’ for any components, please give your reasoning (use the component’s letter to identify which component/s you are referring to):<br><br>Please type as much as you like</td></tr>
<tr><td>2 hours (approximately) to deliver all the session components was:</td><td>Too short ☐</td><td>About right ☐</td><td>Too long ☐</td></tr>
<tr><td colspan="4">Any other comments?<br><br>Please type as much as you like</td></tr>
</table>

WAterS fidelity checklist Session 6 – Allowing feelings

*Please remember:*

- *Complete this ASAP following the session*
- *Fill this out honestly. Remember this is for our learning, not to penalise you*
- *Yes = component fully delivered as per protocol*
- *No = component not delivered or partially delivered*

| Date: | Start time: | End time: |
|---|---|---|
| Name of facilitator/s: | | |
| Number of participants: | | |

| Component | | Delivered as planned? | |
|---|---|---|---|
| Beginning the session (30 mins) | | | |
| A | Re-cap on session 5 | No ☐ | Yes ☐ |
| B | Outline session 6 | No ☐ | Yes ☐ |
| C | Home practice feedback | No ☐ | Yes ☐ |
| Introduction to guided noticing of the breath (15 mins) | | | |
| D | Introduction to guided noticing of the breath | No ☐ | Yes ☐ |
| E | Guided noticing of the breath [audio] | No ☐ | Yes ☐ |
| F | Undertake an enquiry, guided by example questions | No ☐ | Yes ☐ |
| Passengers on the bus after stroke: we all have passengers | | | |
| G | Provide the guided metaphor 'we all have passengers' | No ☐ | Yes ☐ |
| H | Undertake an enquiry, supporting the group to describe, reflect & share | No ☐ | Yes ☐ |
| I | Visual noticing exercise [handbook] | No ☐ | Yes ☐ |
| J | 'What do I tend to do when a passenger shows up' [handbook] | No ☐ | Yes ☐ |
| K | Adequate opportunity for sharing | No ☐ | Yes ☐ |

<table>
<tr><td colspan="4">Ending the session</td></tr>
<tr><td>L</td><td>Summary of the session</td><td>No ☐</td><td>Yes ☐</td></tr>
<tr><td>M</td><td>Adequate opportunity for reflections & comments</td><td>No ☐</td><td>Yes ☐</td></tr>
<tr><td>N</td><td>Home practice [handbook]</td><td>No ☐</td><td>Yes ☐</td></tr>
<tr><td colspan="4">If you’ve answered ‘No’ for any components, please give your reasoning (use the component’s letter to identify which component/s you are referring to):<br><br>Please type as much as you like</td></tr>
</table>

| 2 hours (approximately) to deliver all the session components was: | Too short ☐ | About right ☐ | Too long ☐ |
|---|---|---|---|
| Any other comments?<br><br>*Please type as much as you like* | | | |

WAterS fidelity checklist Session 7 – Making room for feelings

*Please remember:*

- *Complete this ASAP following the session*
- *Fill this out honestly. Remember this is for our learning, not to penalise you*
- *Yes = component fully delivered as per protocol*
- *No = component not delivered or partially delivered*

| Date: | Start time: | End time: |
|---|---|---|
| Name of facilitator/s: | | |
| Number of participants: | | |

| Component | | Delivered as planned? | |
|---|---|---|---|
| Beginning the session (25 mins) | | | |
| A | Re-cap on session 6 | No ☐ | Yes ☐ |
| B | Outline session 7 | No ☐ | Yes ☐ |
| C | Home practice feedback | No ☐ | Yes ☐ |
| Guided mindful noticing of the breath (10 mins) | | | |
| D | Lead guided noticing of the breath practice | No ☐ | Yes ☐ |
| E | Undertake an enquiry, guided by example questions | No ☐ | Yes ☐ |
| Passengers on the bus after stroke: Making room for passengers (25 mins) | | | |
| F | Introduce and provide the guided metaphor 'making room for passengers' | No ☐ | Yes ☐ |
| G | Undertake enquiry, supporting the group to describe, reflect & share | No ☐ | Yes ☐ |
| Walking in the rain metaphor (30 mins) | | | |
| H | Read 'walking in the rain' metaphor | No ☐ | Yes ☐ |
| I | Undertake an enquiry, support the group to reflect & summarise the metaphor | No ☐ | Yes ☐ |
| J | Consolidation & commitments activity [handbook] | No ☐ | Yes ☐ |

| Ending the session (20 mins) | | | |
|---|---|---|---|
| K | Summary of the session | No ☐ | Yes ☐ |
| L | Adequate opportunity for reflections & comments | No ☐ | Yes ☐ |
| M | Home practice [handbook] | No ☐ | Yes ☐ |

If you've answered 'No' for any components, please give your reasoning (use the component's letter to identify which component/s you are referring to):

*Please type as much as you like*

| 2 hours (approximately) to deliver all the session components was: | Too short ☐ | About right ☐ | Too long ☐ |
|---|---|---|---|

Any other comments?

*Please type as much as you like*

WAterS fidelity checklist Session 8 – Doing what it takes

*Please remember:*

- *Complete this ASAP following the session*
- *Fill this out honestly. Remember this is for our learning, not to penalise you*
- *Yes = component fully delivered as per protocol*
- *No = component not delivered or partially delivered*

| Date: | Start time: | End time: |
|---|---|---|
| Name of facilitator/s: | | |
| Number of participants: | | |

| Component | | Delivered as planned? | |
|---|---|---|---|
| Beginning the session (20 mins) | | | |
| A | Re-cap on session 7 | No ☐ | Yes ☐ |
| B | Outline session 8 | No ☐ | Yes ☐ |
| C | Home practice feedback | No ☐ | Yes ☐ |
| Guided mindful colouring practice (15 mins) | | | |
| D | Introduction [handbook] | No ☐ | Yes ☐ |
| E | Guide the mindful colouring practice [handbook & pencils] | No ☐ | Yes ☐ |
| F | Undertake an enquiry, supporting the group to share & reflect | No ☐ | Yes ☐ |
| Stepping stones towards helpful habits (15 mins) | | | |
| G | Stepping stones towards helpful habits [slides] | No ☐ | Yes ☐ |
| H | Undertake an enquiry, supporting the group to share & reflect | No ☐ | Yes ☐ |
| Doing what is takes after stroke plan (25 mins) | | | |
| I | Introduce and explain the doing what it takes after stroke plan [handbook] | No ☐ | Yes ☐ |
| J | Support group to share comments & reflections | No ☐ | Yes ☐ |
| Precious moments: committed action right now (10 mins) | | | |

| | | | |
|---|---|---|---|
| K | Introduce and guide the practice ‘precious moments’ | No ☐ | Yes ☐ |
| L | Undertake a brief enquiry, supporting the group to share & reflect | No ☐ | Yes ☐ |
| Ending the session (20 mins) | | | |
| M | Summary of the session | No ☐ | Yes ☐ |
| N | Adequate opportunity for reflections & comments | No ☐ | Yes ☐ |
| O | Home practice [handbook] | No ☐ | Yes ☐ |
| If you’ve answered ‘No’ for any components, please give your reasoning (use the component’s letter to identify which component/s you are referring to):<br><br>*Please type as much as you like* | | | |
| 2 hours (approximately) to deliver all the session components was: | Too short ☐ | About right ☐ | Too long ☐ |
| Any other comments?<br><br>*Please type as much as you like* | | | |

WaterS fidelity checklist Session 9 – Living better after stroke

*Please remember:*

- *Complete this ASAP following the session*
- *Fill this out honestly. Remember this is for our learning, not to penalise you*
- *Yes = component fully delivered as per protocol*
- *No = component not delivered or partially delivered*

| Date: | Start time: | End time: |
|---|---|---|
| Name of facilitator/s: | | |
| Number of participants: | | |

| | Component | Delivered as planned? | |
|---|---|---|---|
| Beginning the session (20 mins) | | | |
| A | Re-cap on session 8 | No ☐ | Yes ☐ |
| B | Outline session 9 | No ☐ | Yes ☐ |
| C | Home practice feedback | No ☐ | Yes ☐ |
| Guided mindful noticing of the breath (10 mins) | | | |
| D | Lead guided noticing of the breath practice | No ☐ | Yes ☐ |
| E | Undertake a brief enquiry, supporting the group to share & reflect | No ☐ | Yes ☐ |
| Doing what is takes after stroke plan (30 mins) | | | |
| F | Re-introduce the doing what it takes after stroke plan [handbook] | No ☐ | Yes ☐ |
| G | Provide support and facilitation in completion of the plan | No ☐ | Yes ☐ |
| H | Undertake an enquiry, supporting the group to share & reflect | No ☐ | Yes ☐ |
| Ending the session (45 mins) | | | |
| I | Summary of the session and overall course | No ☐ | Yes ☐ |
| J | Adequate opportunity for reflections & comments | No ☐ | Yes ☐ |
| K | Home practice [handbook] | No ☐ | Yes ☐ |

<table>
<tr><td>L</td><td colspan="3">Acknowledge ending of the course and signpost to resources [handbook]</td><td>No ☐</td><td>Yes ☐</td></tr>
<tr><td colspan="6">If you’ve answered ‘No’ for any components, please give your reasoning (use the component’s letter to identify which component/s you are referring to):<br><br>Please type as much as you like</td></tr>
<tr><td colspan="2">2 hours (approximately) to deliver all the session components was:</td><td>Too short ☐</td><td>About right ☐</td><td colspan="2">Too long ☐</td></tr>
<tr><td colspan="6">Any other comments?<br><br>Please type as much as you like</td></tr>
</table>
